# Low lymphocyte count is a risk factor for Parkinson’s disease

**DOI:** 10.1101/2020.09.13.20189530

**Authors:** Melanie P Jensen, Benjamin Meir Jacobs, Ruth Dobson, Sara Bandres-Ciga, Cornelis Blauwendraat, Anette Schrag, The International Parkinson’s Disease Genomics Consortium (IPDGC), Alastair J Noyce

## Abstract

**Importance:** Biomarkers for the early detection of Parkinson’s disease (PD) are needed. Patients with PD display differences in peripheral blood biomarkers of immune function, including leukocyte differential counts and C-reactive protein (CRP), compared to controls. These differences may be useful biomarkers to predict PD, and may shed light on PD pathogenesis.

**Objectives:** To identify whether peripheral immune dysregulation is a pre-diagnostic feature of PD, and whether it plays a causal role.

**Design:** Cross-sectional association analysis of the relationship between differential leukocyte count and other markers of acute inflammation at enrolment, and incident cases of PD in UK Biobank. We used Mendelian randomization to establish whether differences in leukocyte differential counts have a causal influence on risk of PD.

**Setting:** UK Biobank; a population-based cohort with over 500,000 participants aged 40-69 recruited in the UK between 2006 and 2010.

**Participants:** PD cases were defined as individuals with an ICD-10 coded diagnosis of PD. Cases were defined as ’incident’ if their age at diagnosis was greater than their age at recruitment to UKB. ’Controls’ were defined as individuals without a diagnosis of PD. After applying exclusion criteria for pre-existing health conditions that can influence blood counts, 507 incident PD cases and 328,280 controls were included in the analysis.

**Exposure:** Blood cell markers (absolute and relative counts) and other markers of inflammation were obtained from blood tests of participants taken at the initial visit.

**Results:** Lower lymphocyte count was associated with increased odds of incident PD (odds ratio [OR] 0.77, 95% confidence interval [CI] 0.65-0.90). There was weaker evidence of association between lower eosinophil and monocyte counts, lower CRP, and higher neutrophil counts on risk of incident PD. The association between lymphopenia and incident PD remained robust to sensitivity analyses. Mendelian randomization analyses suggested that the effect of low lymphocyte count on PD risk was causal (OR 0.91, 95% CI 0.85 - 0.99).

**Conclusions and relevance:** In this large, prospective setting, lower lymphocyte count was associated with higher risk of subsequent PD diagnosis. Furthermore genetic evidence supported a causal role for lymphocyte count on PD risk.

**Key points:** *Question:* Is the leukcoyte differential count a feature of pre-diagnostic Parkinson’s disease?

*Findings:* In the UK Biobank, a longitudinal cohort study with over 500,000 participants, lower lymphocyte count was associated with a 23% increased odds of incident PD, a significant difference. Mendelian randomisation revealed a convincing causal effect for low lymphocyte count on PD risk.

*Meaning:* Pre-diagnostic Parkinson’s disease is associated with lower lymphocyte counts; the suggestion of causal effect may shed light on PD pathogenesis.

## Introduction

Parkinson’s disease (PD) affects 2% of the population over 65.^1^ The diagnosis is made once motor signs appear, however by this stage ~50% of nigrostriatal neurons have been lost.^2^ There is an urgent unmet clinical need for earlier identification of PD and development of therapies which could slow, prevent, or reverse the progression of the disease.

Immune dysregulation may play a key role in the pathogenesis of PD. The white blood cell (WBC) differential is a crude marker of immune function but is amenable to explore in large-scale observational studies. Studies reveal lower lymphocyte counts in PD patients compared with controls, driven by reductions in helper-CD4+, rather than cytotoxic-CD8+, T-cell and B-cells counts; hypothesized to represent a cytotoxic immune signature.^3-7^ Case-control studies have also identified higher neutrophil and lower lymphocyte counts in patients with established PD compared with controls.^8^

Genetic, epidemiological, and cytokine profiling studies have refined this area of study.^9^ Human Leukocyte Antigen variants (HLA-DRB1/DRB5) have been identified as risk loci for PD in genome-wide association studies (GWAS).^10,11^ Large-scale polygenic risk score analyses suggest this pathway contributes to PD heritability.^12^ *In vitro*, alpha-synuclein-derived peptides are preferentially displayed on major histocompatibility (MHC) molecules associated with PD risk, driving proinflammatory T-cell responses.^13,14^ Variants in the leucine-rich repeat kinase 2 *(LRRK2)* gene, a target for proinflammatory signals, confer effects in the same direction on risk for PD and Crohn’s disease, suggesting common genetic links.^15^ Observational studies have reported reduced risk of PD and reduced penetrance in LRRK2-associated PD with use of immunosuppressants and non-steroidal anti-inflammatory drugs.^16,17^ The prospective ICICLE-PD cohort study found that a baseline ’pro-inflammatory’ cytokine serum profile in PD patients was associated with faster motor deterioration than an ’anti-inflammatory’ profile.^18^

Whether immune dysregulation occurs as an early feature of PD and may be a source of biomarkers, or immune dysregulation has a causal role in driving PD initiation and progression, remains unclear. We studied the relationship between differential leukocyte count and biochemical markers of acute inflammation at enrolment, and incident cases of PD in the UK Biobank (UKB) (https://www.ukbiobank.ac.uk), a large longitudinal cohort with ~500,000 participants. We followed-up signals detected to determine whether differences in leukocyte subsets exerted a causal influence on PD risk using Mendelian randomization (MR).

## Methods

### Population

UKB recruited ~500,000 individuals aged 40-69 between 2006-2010; prospective follow-up data, including census data, blood tests, and healthcare records, are regularly obtained.^19^

PD cases were defined as individuals with an ICD-10 diagnosis of PD (code G20) derived from Hospital Episode Statistics or a self-reported diagnosis of PD. Date at PD diagnosis was determined using the UKB data field ’Date of Parkinson’s Disease report’. Age at diagnosis was derived using this field, age at recruitment, and birth year. Cases were defined as ’incident’ if their age at diagnosis was greater than at recruitment. ’Prevalent’ PD cases, i.e. with a diagnosis of PD at baseline, were excluded from analyses. ’Controls’ were defined as all other individuals in the dataset after applying these exclusions.

Various pre-existing health conditions can influence blood counts. To minimize bias from unbalanced comorbidities among cases and controls we excluded individuals with ICD-10 diagnoses of malignant neoplasms, disease of the blood and blood-forming organs, autoimmune disease, thyrotoxicosis, demyelinating disease of the central nervous system, inflammatory respiratory conditions (asthma and bronchiectasis), non-infective enteritis, inflammatory dermatological conditions (atopic dermatitis and psoriasis), inflammatory polyarthropathies, spondylopathies, and eating disorders (supplementary table 1).

### Blood cell markers

Blood cell markers (absolute and relative counts) and other markers of inflammation (CRP and albumin) were obtained from baseline blood tests of UKB participants taken at the initial assessment visit. Details of data processing can be found on the UKB website (http://biobank.ndph.ox.ac.uk/showcase/showcase/docs/haematology.pdf).

### Statistical analysis

We determined associations of blood cell and inflammatory markers with incident risk of PD using logistic regression. As our primary analysis, we conducted multivariable logistic regression, modelling incident PD diagnosis as the outcome and adjusting for age, sex, Tonwsend deprivation score, and ethnicity (dichotomized as ’White’ background vs all other ethnicities). Models were of the form: Incident PD~Age+Sex+Deprivation+Ethnicity+blood cell marker. The strength of association was determined using the likelihood ratio test, comparing the full model to a null model consisting of the confounding covariates only (Incident PD~Age+Sex+Deprivation+Ethnicity).

We then undertook a variety of sensitivity analyses. First, we included additional covariates in the models: body mass index (BMI) at recruitment, smoking status (“ever” vs “never”), and alcohol consumption (“ever” vs “never”). Second, we excluded individuals within serial time windows of PD diagnosis (<1, <2, <3 years from diagnosis etc.) to determine whether the effects from the primary analysis were restricted to individuals who would go on to develop PD sooner. Third, we repeated the analysis in a matched case:control analysis, individually matching controls by age and sex to PD cases 4:1.

To determine associations between blood markers and time until PD diagnosis, we constructed linear models for the inverse-normal-transformed time to PD diagnosis on age, sex, Townsend score, ethnicity, and blood cell marker. Model fit was quantified using the likelihood ratio test.

### Mendelian randomization

We used MR to determine whether the observational association between lower lymphocyte count and incident PD represents a causal relationship. For MR, SNPs associated with the exposure of interest are used as instrumental variables to estimate the causal effect of the exposure on the outcome.^20,21^ The random allocation of genetic variants from parent to offspring and lifelong exposure mean there are advantages over traditional observational studies in reducing confounding and reverse causation.^22^ We performed two-sample MR using the TwoSampleMR R package.^23,24^ For the exposure instrument, we used summary statistics from the largest published GWAS on blood cell traits from ftp://ftp.sanger.ac.uk/pub/project/humgen/summary_statistics/UKBB_blood_cell_traits/.^25^ This GWAS of 408,112 European UK Biobank participants used as its outcome measure the absolute lymphocyte count adjusted by confounding covariates (age, sex, principal components, study-specific factors), and rank inverse-normalised. Thus a one unit increase in the beta coefficient represents a 1 standard deviation increase in adjusted and normalised lymphocyte count per additional effect allele.

We applied the following to develop a genetic instrument for lymphocyte count:

1. Removed SNPs not typed/imputed in the outcome GWAS;
2. Restricted to biallelic single nucleotide variants;
3. Removed SNPs within the super-extended MHC region due to the complex pleiotropy of this region (hg19, chr6:25,000,000-35,000,000);
4. Restricted to SNPs strongly associated with standardized lymphocyte count (p-value_adjusted_<5e-08);
5. Clumped SNPs using stringent default parameters (LD window=10,000 kb, r^2^=0.001).
6. Removed SNPs explaining more variance in the outcome than the exposure through Steiger filtering.^26^

Outcome data for PD were taken from the most recent and largest case-control GWAS of PD published by the IPDGC and 23andMe.^27^ Overlap in the controls in the exposure and outcome data can result in bias, so we used summary statistics from the PD GWAS that excluded participants from UKB.

We harmonized exposure and outcome SNPs to ensure effect estimates were aligned for the same effect allele. As our primary analysis, we used the inverse-variance weighted (IVW) MR estimate, which provides an accurate causal estimate when MR assumptions are valid.^28^ As secondary sensitivity analyses, we applied the Mixture of Experts approach, which applies different MR estimators and methods for SNP instrument selection (heterogeneity and directionality filtering), and predicts which method has the highest probability of accurately estimating the true causal effect based on the data characteristics.^23^ Power calculations were performed using Stephen Burgess’ online calculator (https://sb452.shinyapps.io/power/).

### Data and code availability

UK Biobank data are available via application (https://www.ukbiobank.ac.uk/). Code is available at https://github.com/benjacobs123456/PD_FBC_UKB. PD GWAS summary statistics which exclude UKB are from Nalls *et al*. 2019 and an application to 23andMe https://research.23andme.com/dataset-access/.^27^ Blood cell trait GWAS summary statistics have been made publicly available by the authors at ftp://ftp.sanger.ac.uk/pub/project/humgen/summary_statistics/UKBB_blood_cell_traits/.

## Results

### Demographics

After applying the exclusion criteria, 507 incident PD cases and 328,280 controls remained in the main (unmatched) analysis. Participant demographic data are shown in table 1.

### Association of blood cell and inflammatory traits with incident PD

In a multivariable logistic regression model adjusting for age, sex, deprivation score, and ethnicity, there was evidence of association (false discovery rate [FDR] Q<0.05) between lower lymphocyte count and incident PD (odds ratio [OR] 0.77, 95% confidence interval [CI] 0.65-0.90, table 2, figure 1). There was evidence of association (FDR Q<0.05) between lower eosinophil count and incident PD but with wide CIs (OR 0.33, 95% CI 0.14-0.76, table 2). There was weaker evidence (FDR Q<0.10) of associations between lower CRP, lower monocyte count, higher neutrophil count and increased risk of incident PD (table 2).

**Figure 1:**
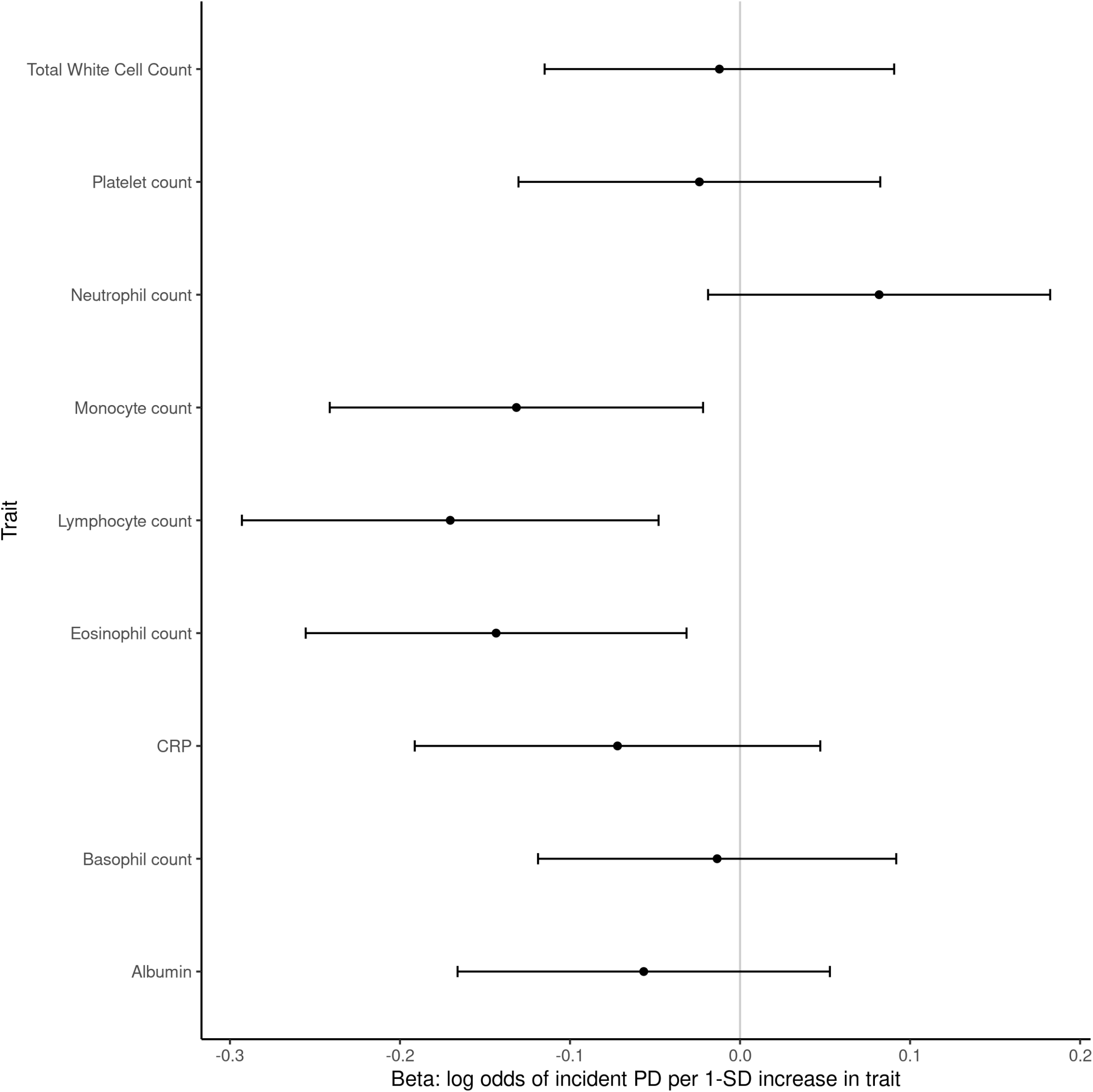
Association of blood cell marker and inflammatory markers with incident PD in UKB. Betas and 95% CIs are shown from multivariable logistic regression models of the form PD ~ Age + Sex + Ethnicity + Townsend deprivation score + blood biomarker. Estimates shown here are for Z-score standardised variables (x-mean / sd) to allow comparison between effect sizes.

To determine whether these associations could be driven by confounding, we constructed models in which we also controlled for variables which can impact both PD risk and blood cell indices: BMI, smoking, and alcohol consumption. The effect estimates from these models were less precise but of a similar magnitude to the primary analysis (lymphocyte count OR 0.78, 95% CI 0.66-0.92; eosinophil count OR 0.38, 95% CI 0.16-0.87; CRP OR 0.97, 95% CI 0.94-1.01; monocyte count OR 0.60, 95% CI 0.34-1.06; neutrophil account OR 1.08, 95% CI 1.01-1.15 on risk of incident PD) (supplementary table 2).

To examine the possibility that reverse causation could be driving our findings - i.e. that early PD could be driving lower lymphocyte counts -we excluded individuals who underwent blood draw within 5 years of PD diagnosis. Despite the loss of power resulting from this restriction, we obtained similar effect estimates (supplementary table 3). To examine whether the association between lower lymphocyte count and higher PD risk persisted through the pre-diagnostic period, we repeated this analysis excluding individuals within serial time windows of diagnosis (within 1, 2, 3, 4, 5, 6, 7, and 8 years of diagnosis). Despite lower numbers of cases with increasing time pre-diagnosis, there was a consistent signal in all groups (supplementary table 4). We obtained similar results for the association between lower lymphocyte count and risk of incident PD with the matched case-control analysis (OR 0.78, 95% CI 0.65-0.93) (supplementary table 5). Lymphopenia considered as a binary trait (defined as absolute lymphocyte count<1×10^9^ cells/L), was strongly associated with a higher risk of PD (OR 1.93, 95% CI 1.26-2.97, p=0.006). Exclusion of extreme lymphocyte counts (mean ± >3SD, leaving an inclusion lymphocyte count range 0.05-3.85×10^9^ cells/L) did not substantially alter the observed association (OR 0.76 per 1-SD increase in lymphocyte count, 95% CI 0.64-0.90).

To assess whether inflammatory and blood cell markers were associated with time until PD diagnosis, we constructed linear models adjusting for age, sex, deprivation score and ethnicity. There was no strong evidence that any blood markers were associated with time until PD diagnosis (FDR >0.05, supplementary table 6).

### Mendelian randomization

To generate an instrument for lymphocyte count, we excluded SNPs not typed/imputed in both the exposure and outcome datasets, SNPs with p≥5e^-08^, SNPs lying within the super-extended MHC (chr6:25,000,000-35,000,000 on genome build hg19), and palindromic SNPs with intermediate effect allele frequencies. We performed LD clumping using default parameters and Steiger directionality filtering, yielding a genetic instrument of 510 independent non-MHC autosomal SNPs associated with lymphocyte count (supplementary table 7). Collectively these SNPs explained 5.37% of the variance in lymphocyte count in this sample and could be considered a strong instrument (F statistic=45.3).

The primary MR analysis (IVW) showed evidence of a causal effect of lymphocyte count on PD risk commensurate with the size of the observational effect (OR 0.91, 95% CI 0.85-0.99, p=0.023). There was no evidence that unbalanced horizontal pleiotropy (whereby variants influence the outcome via pathways other than through the exposure) was biasing the IVW result (MR-Egger intercept 0.0006, p=0.76). There was evidence of substantial heterogeneity in the IVW estimate (Cochran’s Q=642, p=5.29e^-05^), however heterogeneity filtering did not alter the magnitude of the effect (OR 0.91, 95% CI 0.85-0.97, p=0.0034), and in fact increased the precision of the estimate. Standard sensitivity analyses and the ’mixture of experts’ approach yielded similar causal effect estimates with varying degrees of precision, collectively providing evidence of a causal effect of genetically-determined lymphocyte count on PD risk (figure 2, supplementary table 8).

**Figure 2:**
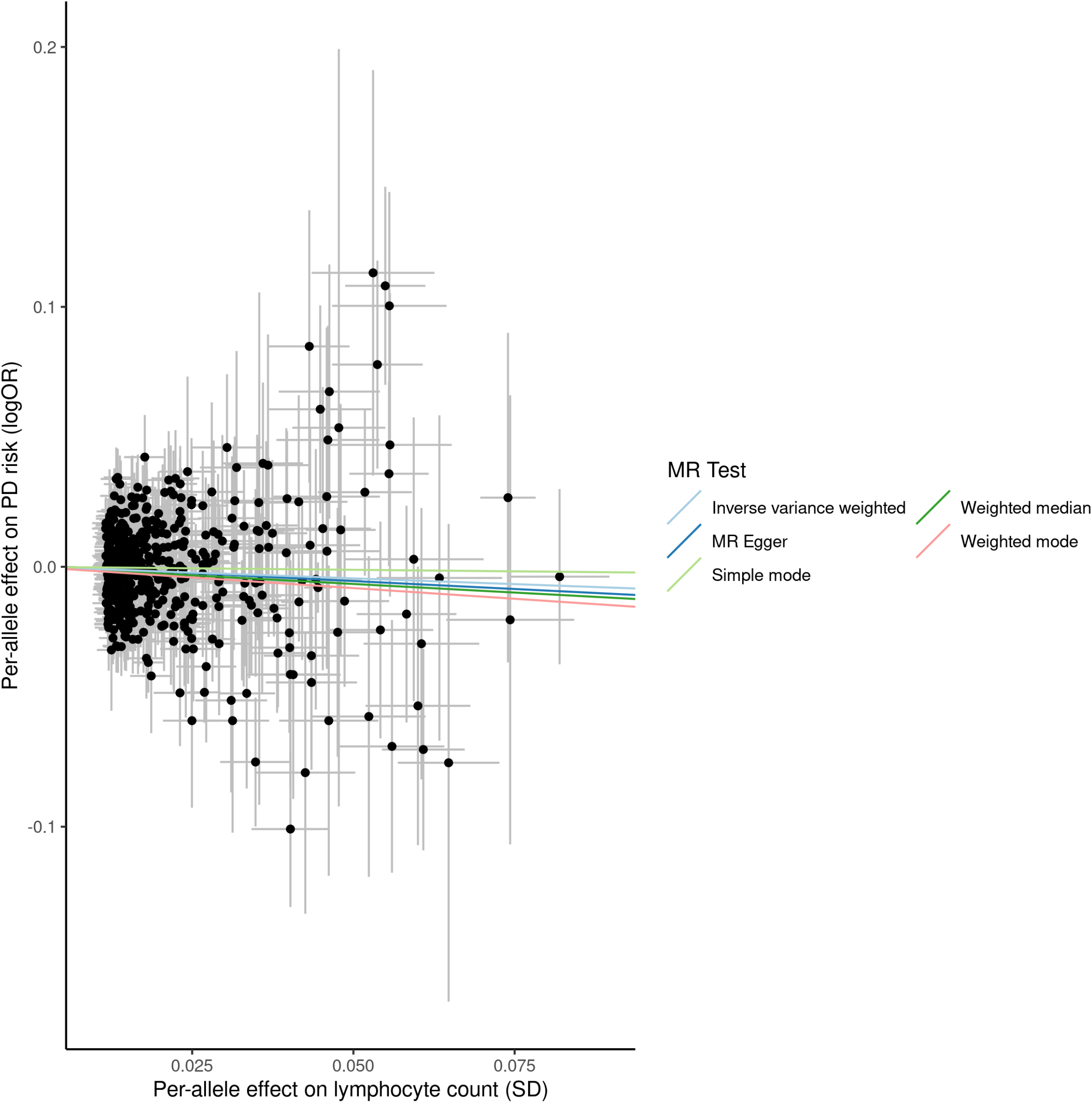
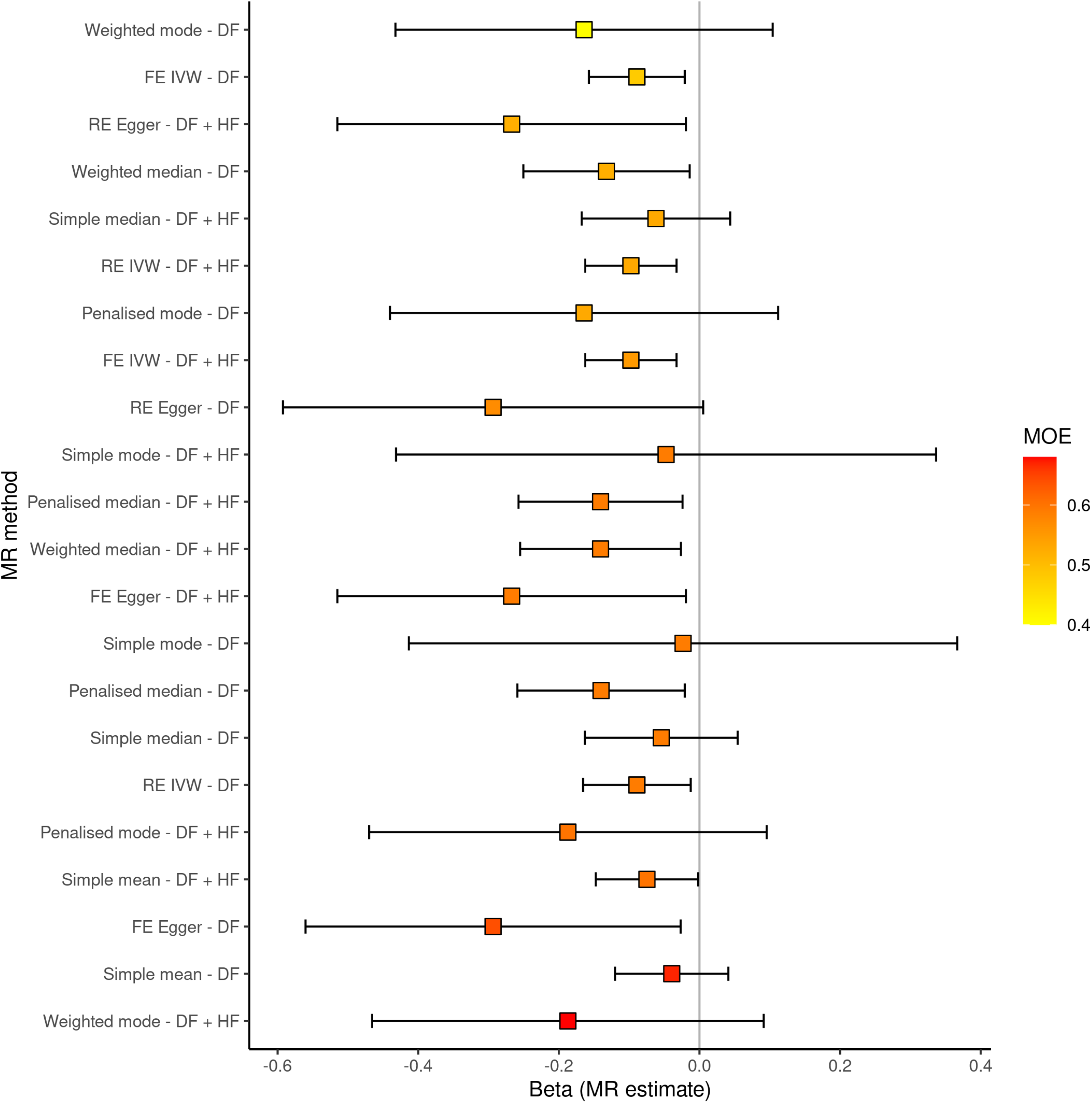
Mendelian randomisation analysis of the effect of lymphocyte count on PD risk. A: scatter plot showing SNP associations with lymphocyte count from our GWAS (x) and with the per-allele log(OR) for PD (estimates from the largest PD GWAS published, Meta5, excluding UKB participants). The model fit lines indicate MR estimates (of the ’causal effect) from different MR methods. B: MR estimates from various methods using the ’Mixture of Experts approach’. The y axis shows different MR methods and different approaches for filtering SNPs to be included in the genetic instrument (heterogeneity filtering, HF; directionality filtering, DF). Estimates are coloured and ordered by the ’MOE’ statistic, which is similar to an area under the curve statistic in that it quantifies that ability of a given MR method to distinguish a true effect from the absence of a true effect. MOE statistics closer to 1 indicate a higher likelihood that the given MR method will give an accurate estimate for the given dataset.

## Discussion

In this prospective cohort involving >320 000 individuals, lower lymphocyte count was associated with increased risk of incident PD; marginal associations were observed for lower eosinophil and monocyte counts, and higher neutrophil counts. The association between lymphopenia and incident PD risk remained robust to several sensitivity analyses. Follow-up MR analyses revealed evidence supporting the hypothesis that lower lymphocyte count may be a causal risk factor for developing PD later in life.

Only one study has explored the link between leukocyte subsets and risk of incident PD. In the Swedish Apolipoprotein-Related Mortality Risk cohort, lower lymphocyte count was associated with a lower risk of incident PD (HR 0.74, 95% CI 0.59-0.94).^29^ However, pre-existing health conditions which can influence blood counts were not excluded or adjusted for so the possibility of residual confounding remained; we applied stringent exclusion criteria capturing comorbidities which influence leukocyte counts, such as autoimmune disease and cancer.

Several studies have assessed changes in leukocyte populations in PD after diagnosis. Established PD is associated with lymphopenia, driven by absolute reductions in CD4^+^ T-helper cells, CD19^+^ B-cells and Treg cells.^4,5,30,31^ In a study of 123 newly diagnosed PD patients, the percentage of neutrophils and lymphocytes had positive and negative correlations, respectively, with UPDRS motor scores.^32^ This has been interpreted as evidence to support an autoimmune/inflammatory component in PD pathogenesis.^33^

Reductions in the absolute number of naïve-T/B-cells is also a feature of age-related immune dysregulation, leading to the hypothesis that PD is a state of premature ageing.^34-36^ Flow cytometric analysis has demonstrated that leukocyte apoptosis is higher in PD patients than controls and is associated with dopaminergic deficits on SPECT.^37^ However, studies to date have examined immune markers in established PD and cannot determine whether dysregulated adaptive immunity causally influences PD risk or represents a downstream consequence of PD pathology. Moreover, most PD cases in these studies are receiving dopaminergic medication, which could confound immunophenotypic patient/control differences.^38-40^ We demonstrate that lower lymphocyte count may be a feature of PD at least 8 years before diagnosis, before the initiation of dopaminergic therapy, extending the observations made in patients with established PD into the pre-diagnostic phase.

The observed association between lymphopenia and incident PD may be driven by a causal effect, residual confounding or reverse causation. Our MR analyses suggest a causative effect of low lymphocyte count on PD risk. Specifically, the IVW estimate suggests a weak causal effect of lymphocyte count on PD risk such that each 1-SD increase in adjusted lymphocyte count decreased the odds of PD by 9%. The MR-Egger regression intercept, which quantifies the degree to which net unbalanced horizontal pleiotropy may bias the IVW estimate, was close to null, suggesting that the IVW estimate provides a robust estimate of the true causal effect. Sensitivity analyses using a variety of MR estimators and heterogeneity filtering supported the magnitude of the IVW estimate, albeit with varying precision. These data lend weight to the view that low lymphocyte count is not purely an effect of PD and its prodromes (reverse causation) or confounding, but may be involved in a causal chain of events contributing to the development of PD.

To address confounding in the observational study, we corrected for potential confounders in the primary analysis and undertook sensitivity analyses (further addition of covariates and a matched analysis). However residual confounding, that is pre-diagnostic PD affects lymphocyte count, may remain. Although the effect persisted after excluding individuals who underwent blood draw within 8 years of diagnosis, pre-diagnostic features of PD including constipation may predate clinical diagnosis by 20 years.^41,42^ The gut has been proposed as the site of initiation of PD and fecal microbiome changes are also noted in prodromal PD.^43^ In turn, manipulation of intestinal microbiota in mice has been shown to directly modulate lymphopenia-induced autoimmunity.^44^ Lymphopenia may be a feature of pre-diagnostic PD, and reflect early pathological changes in peripheral tissues.

Higher CRP was marginally associated with reduced risk of PD. A meta-analysis of 23 case-control studies found significantly higher levels of CRP in the serum, cerebrospinal (CSF), and whole blood of manifest PD subjects compared with controls.^45^ CSF CRP levels tend to increase over time from PD diagnosis and the development of PD-dementia.^46,47^ These findings suggest that systemic inflammation impacts or occurs as a consequence of disease progression, but does not shed light on whether it influences disease initiation.^9^

Strengths of our study include the large sample size derived from UKB. Despite low response rates potentially leading to more favorable risk factor profiles, exposure-outcome associations in the UKB seem to be generalisable.^48^ The comprehensive phenotyping of individuals in the cohort allowed us to correct for multiple potential confounding factors. In contrast to previous work utilizing cohorts with manifest PD, the availability of baseline blood tests and longitudinal assessment of PD diagnosis enabled us to examine the association between leukocyte subsets and risk of incident PD.

Limitations of our study include the lack of adjustment for medication which could impact leukocyte subsets, although exclusion for comorbidity will have captured a significant proportion of this confounding. Due to lack of flow cytometric data in UKB, we could not establish whether the observed association was driven by reductions in T-cells and/or B-cells. MR analysis precludes the identification of non-linear exposure-outcome associations, although non-linear mechanisms are not seen in other conditions in which lymphopenia influences outcome.^49,50^

In conclusion we report that lower lymphocyte count was associated with higher risk of subsequent diagnosis of PD in a large UK cohort. The association remained robust to a range of sensitivity analyses and persisted at 8 years pre-diagnosis. MR analyses suggested that this relationship may be causal and this in particular warrants further exploration. It is also plausible that lymphopenia may be a consequence of pre-diagnostic PD which, although lacking specificity in isolation, could enhance efforts to identify those in the earliest stages of PD.^51^ Further work is required to replicate these findings in other cohorts, address the mechanisms by which lymphopenia may causally intersect with the pathobiology of PD, and evaluate whether lymphopenia improves algorithms aiming to predict who will develop PD in prospective cohort designs.

## Data Availability

UK Biobank data are available via application (https://www.ukbiobank.ac.uk/). Code is available at https://github.com/benjacobs123456/PD_FBC_UKB. PD GWAS summary statistics which exclude UKB are from Nalls et al. 2019 and an application to 23andMe https://research.23andme.com/dataset-access/. Blood cell trait GWAS summary statistics have been made publicly available by the authors at ftp://ftp.sanger.ac.uk/pub/project/humgen/summary_statistics/UKBB_blood_cell_traits/.

## Acknowledgements

This research has been conducted using the UK Biobank Resource. We thank the participants of the UK Biobank. We would also like to acknowledge the 23andMe Research Team (especially Dr Karl Heilbron) and the 23andMe research participants.We acknowledge the IPDGC consortium members and affiliations:

<u>**United Kingdom:**</u>

<u>Alastair J Noyce</u> (Preventive Neurology Unit, Wolfson Institute of Preventive Medicine, QMUL, London, UK and Department of Molecular Neuroscience, UCL, London, UK), <u>Rauan Kaiyrzhanov</u> (Department of Molecular Neuroscience, UCL Institute of Neurology, London, UK), <u>Ben Middlehurst</u> (Institute of Translational Medicine, University of Liverpool, Liverpool, UK), <u>Demis A Kia</u> (UCL Genetics Institute; and Department of Molecular Neuroscience, UCL Institute of Neurology, London, UK), <u>Manuela Tan</u> (Department of Clinical Neuroscience, University College London, London, UK), <u>Henry Houlden</u> (Department of Molecular Neuroscience, UCL Institute of Neurology, London, UK), <u>Catherine S Storm</u> (Department of Clinical and Movement Neurosciences, UCL Queen Square Institute of Neurology, London, UK), <u>Huw R Morris</u> (Department of Clinical Neuroscience, University College London, London, UK), <u>Helene Plun-Favreau</u> (Department of Molecular Neuroscience, UCL Institute of Neurology, London, UK), <u>Peter Holmans</u> (Biostatistics & Bioinformatics Unit, Institute of Psychological Medicine and Clinical Neuroscience, MRC Centre for Neuropsychiatric Genetics & Genomics, Cardiff, UK), <u>John Hardy</u> (Department of Molecular Neuroscience, UCL Institute of Neurology, London, UK), <u>Daniah Trabzuni</u> (Department of Molecular Neuroscience, UCL Institute of Neurology, London, UK; Department of Genetics, King Faisal Specialist Hospital and Research Centre, Riyadh, 11211 Saudi Arabia), <u>John Quinn</u> (Institute of Translational Medicine, University of Liverpool, Liverpool, UK), <u>Vivien Bubb</u> (Institute of Translational Medicine, University of Liverpool, Liverpool, UK), <u>Kin Y Mok</u> (Department of Molecular Neuroscience, UCL Institute of Neurology, London, UK), <u>Kerri J. Kinghorn</u> (Institute of Healthy Ageing, Research Department of Genetics, Evolution and Environment, University College London, London, UK), <u>Nicholas W Wood</u> (UCL Genetics Institute; and Department of Molecular Neuroscience, UCL Institute of Neurology, London, UK), <u>Patrick Lewis</u> (University of Reading, Reading, UK), <u>Sebastian R Schreglmann</u> (Department of Molecular Neuroscience, UCL Institute of Neurology, London, UK), <u>Ruth Lovering</u> (University College London, London, UK), <u>Lea R’Bibo</u> (Department of Molecular Neuroscience, UCL Institute of Neurology, London, UK), <u>Claudia Manzoni</u> (University of Reading, Reading, UK), <u>Mie Rizig</u> (Department of Molecular Neuroscience, UCL Institute of Neurology, London, UK), <u>Mina Ryten</u> (Department of Molecular Neuroscience, UCL Institute of Neurology, London, UK), <u>Sebastian Guelfi</u> (Department of Molecular Neuroscience, UCL Institute of Neurology, London, UK), <u>Valentina Escott-Price</u> (MRC Centre for Neuropsychiatric Genetics and Genomics, Cardiff University School of Medicine, Cardiff, UK), Viorica Chelban (Department of Molecular Neuroscience, UCL Institute of Neurology, London, UK), <u>Thomas Foltynie</u> (UCL Institute of Neurology, London, UK), <u>Nigel Williams</u> (MRC Centre for Neuropsychiatric Genetics and Genomics, Cardiff, UK), Karen E. Morrison (Faculty of Medicine, University of Southampton, UK), Carl Clarke (University of Birmingham, Birmingham, UK and Sandwell and West Birmingham Hospitals NHS Trust, Birmingham, UK), <u>Kirsten Harvey</u> (UCL School of Pharmacy, UK), <u>Benjamin M Jacobs</u> (Preventive Neurology Unit, Wolfson Institute of Preventive Medicine, QMUL, London, UK).

<u>**France:**</u>

<u>Alexis Brice</u> (Institut du Cerveau et de la Moelle épinière, ICM, Inserm U 1127, CNRS, UMR 7225, Sorbonne Universités, UPMC University Paris 06, UMR S 1127, AP-HP, Pitié-Salpêtrière Hospital, Paris, France), <u>Fabrice Danjou</u> (Institut du Cerveau et de la Moelle épinière, ICM, Inserm U 1127, CNRS, UMR 7225, Sorbonne Universités, UPMC University Paris 06, UMR S 1127, AP-HP, Pitié-Salpêtrière Hospital, Paris, France), <u>Suzanne Lesage</u> (Institut du Cerveau et de la Moelle épinière, ICM, Inserm U 1127, CNRS, UMR 7225, Sorbonne Universités, UPMC University Paris 06, UMR S 1127, AP-HP, Pitié-Salpêtrière Hospital, Paris, France), <u>Jean-Christophe Corvol</u> (Institut du Cerveau et de la Moelle épinière, ICM, Inserm U 1127, CNRS, UMR 7225, Sorbonne Universités, UPMC University Paris 06, UMR S 1127, Centre d’Investigation Clinique Pitié Neurosciences CIC-1422, AP-HP, Pitié-Salpêtrière Hospital, Paris, France), <u>Maria Martinez</u> (INSERM UMR 1220; and Paul Sabatier University, Toulouse, France),

<u>**Germany:**</u>

<u>Claudia Schulte</u> (Department for Neurodegenerative Diseases, Hertie Institute for Clinical Brain Research, University of Tübingen, and DZNE, German Center for Neurodegenerative Diseases, Tübingen, Germany), <u>Kathrin Brockmann</u> (Department for Neurodegenerative Diseases, Hertie Institute for Clinical Brain Research, University of Tübingen, and DZNE, German Center for Neurodegenerative Diseases, Tübingen, Germany), <u>Javier Simón-Sánchez</u> (Department for Neurodegenerative Diseases, Hertie Institute for Clinical rain Research, University of Tübingen, and DZNE, German Center for Neurodegenerative Diseases, Tübingen, Germany), <u>Peter Heutink</u> (DZNE, German Center for Neurodegenerative Diseases and Department for Neurodegenerative Diseases, Hertie Institute for Clinical Brain Research, University of Tübingen, Tübingen, Germany), <u>Patrizia Rizzu</u> (DZNE, German Center for Neurodegenerative Diseases), <u>Manu Sharma</u> (Centre for Genetic Epidemiology, Institute for Clinical Epidemiology and Applied Biometry, University of Tubingen, Germany), <u>Thomas Gasser</u> (Department for Neurodegenerative Diseases, Hertie Institute for Clinical Brain Research, and DZNE, German Center for Neurodegenerative Diseases, Tübingen, Germany), <u>Susanne A. Schneider</u> (Department of Neurology, Ludwig-Maximilians-University Munich, München, Germany)

<u>**United States of America:**</u>

<u>Mark R Cookson</u> (Laboratory of Neurogenetics, National Institute on Aging, Bethesda, USA), <u>Sara Bandres-Ciga</u> (Laboratory of Neurogenetics, National Institute on Aging, Bethesda, MD, USA), <u>Cornelis Blauwendraat</u> (Laboratory of Neurogenetics, National Institute on Aging, Bethesda, MD, USA), <u>David W. Craig</u> (Department of Translational Genomics, Keck School of Medicine, University of Southern California, Los Angeles, USA), <u>Kimberley Billingsley</u> (Laboratory of Neurogenetics, National Institute on Aging, Bethesda, MD, USA), Mary B. Makarious (Laboratory of Neurogenetics, National Institute on Aging, Bethesda, MD, USA), <u>Derek P. Narendra</u> (Inherited Movement Disorders Unit, National pInstitute of Neurological Disorders and Stroke, Bethesda, MD, USA), <u>Faraz Faghri</u> (Laboratory of Neurogenetics, National Institute on Aging, Bethesda, USA; Department of Computer Science, University of Illinois at Urbana-Champaign, Urbana, IL, USA), <u>J Raphael Gibbs</u> (Laboratory of Neurogenetics, National Institute on Aging, National Institutes of Health, Bethesda, MD, USA), <u>Dena G. Hernandez</u> (Laboratory of Neurogenetics, National Institute on Aging, Bethesda, MD, USA), <u>Kendall Van Keuren-Jensen</u> (Neurogenomics Division, TGen, Phoenix, AZ USA), <u>Joshua M. Shulman</u> (Departments of Neurology, Neuroscience, and Molecular & Human Genetics, Baylor College of Medicine, Houston, Texas, USA; Jan and Dan Duncan Neurological Research Institute, Texas Children’s Hospital, Houston, Texas, USA), <u>Hirotaka Iwaki</u> (Laboratory of Neurogenetics, National Institute on Aging, Bethesda, MD, USA), <u>Hampton L. Leonard</u> (Laboratory of Neurogenetics, National Institute on Aging, Bethesda, MD, USA), <u>Mike A. Nalls</u> (Laboratory of Neurogenetics, National Institute on Aging, Bethesda, USA; CEO/Consultant Data Tecnica International, Glen Echo, MD, USA), <u>Laurie Robak</u> (Baylor College of Medicine, Houston, Texas, USA), <u>Jose Bras</u> (Center for Neurodegenerative Science, Van Andel Research Institute, Grand Rapids, Michigan, USA), <u>Rita Guerreiro</u> (Center for Neurodegenerative Science, Van Andel Research Institute, Grand Rapids, Michigan, USA), <u>Steven Lubbe</u> (Ken and Ruth Davee Department of Neurology and Simpson Querrey Center for Neurogenetics, Northwestern University Feinberg School of Medicine, Chicago, IL, USA), <u>Steven Finkbeiner</u> (Departments of Neurology and Physiology, University of California, San Francisco; Gladstone Institute of Neurological Disease; Taube/Koret Center for Neurodegenerative Disease Research, San Francisco, CA, USA), <u>Niccolo E. Mencacci</u> (Northwestern University Feinberg School of Medicine, Chicago, IL, USA), <u>Codrin Lungu</u> (National Institutes of Health Division of Clinical Research, NINDS, National Institutes of Health, Bethesda, MD, USA), <u>Andrew B Singleton</u> (Laboratory of Neurogenetics, National Institute on Aging, Bethesda, MD, USA), <u>Sonja W. Scholz</u> (Neurodegenerative Diseases Research Unit, National Institute of Neurological Disorders and Stroke, Bethesda, MD, USA), <u>Xylena Reed</u> (Laboratory of Neurogenetics, National Institute on Aging, Bethesda, MD, USA). <u>Roy N. Alcalay</u> (Department of Neurology, College of Physicians and Surgeons, Columbia University Medical Center, New York, NY, USA, Taub Institute for Research on Alzheimer’s Disease and the Aging Brain, College of Physicians and Surgeons, Columbia University Medical Center, New York, NY, USA). <u>Zbigniew K. Wszolek</u> (Department of Neurology, Mayo Clinic Jacksonville, FL, USA), <u>Ryan J. Uitti</u> (Department of Neurology, Mayo Clinic Jacksonville, FL, USA), <u>Owen A. Ross</u> (Departments of Neuroscience & Clinical Genomics, Mayo Clinic Jacksonville, FL, USA), <u>Francis P. Grenn</u> (Laboratory of Neurogenetics, National Institute on Aging, Bethesda, MD, USA), <u>Anni Moore</u> (Laboratory of Neurogenetics, National Institute on Aging, Bethesda, MD, USA).

<u>**Canada:**</u>

<u>Ziv Gan-Or</u> (Montreal Neurological Institute and Hospital, Department of Neurology & Neurosurgery, Department of Human Genetics, McGill University, Montréal, QC, H3A 0G4, Canada), <u>Guy A. Rouleau</u> (Montreal Neurological Institute and Hospital, Department of Neurology & Neurosurgery, Department of Human Genetics, McGill University, Montréal, QC, H3A 0G4, Canada), <u>Lynne Krohn</u> (Montreal Neurological Institute and Hospital, Department of Neurology & Neurosurgery, Department of Human Genetics, McGill University, Montréal, QC, H3A 0G4, Canada), <u>Kheireddin Mufti</u> (Montreal Neurological Institute and Hospital, Department of Neurology & Neurosurgery, Department of Human Genetics, McGill University, Montréal, QC, H3A 0G4, Canada),

<u>**The Netherlands:**</u>

<u>Jacobus J van Hilten</u> (Department of Neurology, Leiden University Medical Center, Leiden, Netherlands), <u>Johan Marinus</u> (Department of Neurology, Leiden University Medical Center, Leiden, Netherlands)

<u>**Spain:**</u>

<u>Astrid D. Adarmes-Gómez</u> (Instituto de Biomedicina de Sevilla (IBiS), Hospital Universitario Virgen del Rocío/CSIC/Universidad de Sevilla, Seville), <u>Miquel Aguilar</u> (Fundació Docència i Recerca Mútua de Terrassa and Movement Disorders Unit, Department of Neurology, University Hospital Mutua de Terrassa, Terrassa, Barcelona.), <u>Ignacio Alvarez</u> (Fundació Docència i Recerca Mútua de Terrassa and Movement Disorders Unit, Department of Neurology, University Hospital Mutua de Terrassa, Terrassa, Barcelona.),<u>Victoria Alvarez</u> (Hospital Universitario Central de Asturias, Oviedo), <u>Francisco Javier Barrero</u> (Hospital Universitario San Cecilio de Granada, Universidad de Granada), <u>Jesús Alberto Bergareche Yarza</u> (Instituto de Investigación Sanitaria Biodonostia, San Sebastián), <u>Inmaculada Bernal-Bernal</u> (Instituto de Biomedicina de Sevilla (IBiS), Hospital Universitario Virgen del Rocío/CSIC/Universidad de Sevilla, Seville), <u>Marta Blazquez</u> (Hospital Universitario Central de Asturias, Oviedo), <u>Marta Bonilla-Toribio</u> (Instituto de Biomedicina de Sevilla (IBiS), Hospital Universitario Virgen del Rocío/CSIC/Universidad de Sevilla, Seville), <u>Juan A. Botía</u> (Universidad de Murcia, Murcia), <u>María Teresa Boungiorno</u> (Fundació Docència i Recerca Mútua de Terrassa and Movement Disorders Unit, Department of Neurology, University Hospital Mutua de Terrassa, Terrassa, Barcelona.) <u>Dolores Buiza-Rueda</u> (Instituto de Biomedicina de Sevilla (IBiS), Hospital Universitario Virgen del Rocío/CSIC/Universidad de Sevilla, Seville), <u>Ana Cámara</u> (Hospital Clinic de Barcelona), Fátima Carrillo (Instituto de Biomedicina de Sevilla (IBiS), Hospital Universitario Virgen del Rocío/CSIC/Universidad de Sevilla, Seville), <u>Mario Carrión-Claro</u> (Instituto de Biomedicina de Sevilla (IBiS), Hospital Universitario Virgen del Rocío/CSIC/Universidad de Sevilla, Seville), <u>Debora Cerdan</u> (Hospital General de Segovia, Segovia), <u>Jordi Clarimón</u> (Memory Unit, Department of Neurology, IIB Sant Pau, Hospital de la Santa Creu i Sant Pau, Universitat Autònoma de Barcelona and Centro de Investigación Biomédica en Red en Enfermedades Neurodegenerativas (CIBERNED), Madrid),<u>Yaroslau Compta</u> (Hospital Clinic de Barcelona), <u>Monica Diez-Fairen</u> (Fundació Docència i Recerca Mútua de Terrassa and Movement Disorders Unit, Department of Neurology, University Hospital Mutua de Terrassa, Terrassa, Barcelona.), <u>Oriol Dols-Icardo</u> (Memory Unit, Department of Neurology, IIB Sant Pau, Hospital de la Santa Creu i Sant Pau, Universitat Autònoma de Barcelona, Barcelona, and Centro de Investigación Biomédica en Red en Enfermedades Neurodegenerativas (CIBERNED), Madrid), <u>Jacinto Duarte</u> (Hospital General de Segovia, Segovia), <u>Raquel Duran</u> (Centro de Investigacion Biomedica, Universidad de Granada, Granada), <u>Francisco Escamilla-Sevilla</u> (Hospital Universitario Virgen de las Nieves, Instituto de Investigación Biosanitaria de Granada, Granada), <u>Mario Ezquerra</u> (Hospital Clinic de Barcelona), <u>Cici Feliz</u> (Departmento de Neurologia, Instituto de Investigación Sanitaria Fundación Jiménez Díaz, Madrid, Spain), <u>Manel Fernández</u> (Hospital Clinic de Barcelona), <u>Rubén Femández-Santiago</u> (Hospital Clinic de Barcelona), <u>Ciara Garcia</u> (Hospital Universitario Central de Asturias, Oviedo), <u>Pedro García-Ruiz</u> (Instituto de Investigación Sanitaria Fundación Jiménez Díaz, Madrid), <u>Pilar Gómez-Garre</u> (Instituto de Biomedicina de Sevilla (IBiS), Hospital Universitario Virgen del Rocío/CSIC/Universidad de Sevilla, Seville), <u>Maria Jose Gomez Heredia</u> (Hospital Universitario Virgen de la Victoria, Malaga), <u>Isabel Gonzalez-Aramburu</u> (Hospital Universitario Marqués de Valdecilla-IDIVAL, Santander),<u>Ana Gorostidi Pagola</u> (Instituto de Investigación Sanitaria Biodonostia, San Sebastián), <u>Janet Hoenicka</u> (Institut de Recerca Sant Joan de Déu, Barcelona), <u>Jon Infante</u> (Hospital Universitario Marqués de Valdecilla-IDIVAL and University of Cantabria, Santander, and Centro de Investigación Biomédica en Red en Enfermedades Neurodegenerativas (CIBERNED), <u>Silvia Jesús</u> (Instituto de Biomedicina de Sevilla (IBiS), Hospital Universitario Virgen del Rocío/CSIC/Universidad de Sevilla, Seville), <u>Adriano Jimenez-Escrig</u> (Hospital Universitario Ramón y Cajal, Madrid), <u>Jaime Kulisevsky</u> (Movement Disorders Unit, Department of Neurology, IIB Sant Pau, Hospital de la Santa Creu i Sant Pau, Universitat Autònoma de Barcelona, Barcelona, and Centro de Investigación Biomédica en Red en Enfermedades Neurodegenerativas (CIBERNED)), <u>Miguel A. Labrador-Espinosa</u> (Instituto de Biomedicina de Sevilla (IBiS), Hospital Universitario Virgen del Rocío/CSIC/Universidad de Sevilla, Seville), <u>Jose Luis Lopez-Sendon</u> (Hospital Universitario Ramón y Cajal, Madrid), <u>Adolfo López de Munain Arregui</u> (Instituto de Investigación Sanitaria Biodonostia, San Sebastián), <u>Daniel Macias</u> (Instituto de Biomedicina de Sevilla (IBiS), Hospital Universitario Virgen del Rocío/CSIC/Universidad de Sevilla, Seville), <u>Irene Martínez Torres</u> (Department of Neurology, Instituto de Investigación Sanitaria La Fe, Hospital Universitario y Politécnico La Fe, Valencia), <u>Juan Marín</u> (Movement Disorders Unit, Department of Neurology, IIB Sant Pau, Hospital de la Santa Creu i Sant Pau, Universitat Autònoma de Barcelona, Barcelona, and Centro de Investigación Biomédica en Red en Enfermedades Neurodegenerativas (CIBERNED)), <u>Maria Jose Marti</u> (Hospital Clinic Barcelona), <u>Juan Carlos Martínez-Castrillo</u> (Instituto Ramón y Cajal de Investigación Sanitaria, Hospital Universitario Ramón y Cajal, Madrid), <u>Carlota Méndez-del-Barrio</u> (Instituto de Biomedicina de Sevilla (IBiS), Hospital Universitario Virgen del Rocío/CSIC/Universidad de Sevilla, Seville), <u>Manuel Menéndez González</u> (Hospital Universitario Central de Asturias, Oviedo), <u>Marina Mata</u> (Department of Neurology, Hospital Universitario Infanta Sofía, Madrid, Spain) <u>Adolfo Mínguez</u> (Hospital Universitario Virgen de las Nieves, Granada, Instituto de Investigación Biosanitaria de Granada), <u>Pablo Mir</u> (Instituto de Biomedicina de Sevilla (IBiS), Hospital Universitario Virgen del Rocío/CSIC/Universidad de Sevilla, Seville), <u>Elisabet Mondragon Rezola</u> (Instituto de Investigación Sanitaria Biodonostia, San Sebastián), <u>Esteban Muñoz</u> (Hospital Clinic Barcelona), <u>Javier Pagonabarraga</u> (Movement Disorders Unit, Department of Neurology, IIB Sant Pau, Hospital de la Santa Creu i Sant Pau, Universitat Autònoma de Barcelona, Barcelona, and Centro de Investigación Biomédica en Red en Enfermedades Neurodegenerativas (CIBERNED)), <u>Pau Pastor</u> (Fundació Docència i Recerca Mútua de Terrassa and Movement Disorders Unit, Department of Neurology, University Hospital Mutua de Terrassa, Terrassa, Barcelona.), <u>Francisco Perez Errazquin</u> (Hospital Universitario Virgen de la Victoria, Malaga), <u>Teresa Periñán-Tocino</u> (Instituto de Biomedicina de Sevilla (IBiS), Hospital Universitario Virgen del Rocío/CSIC/Universidad de Sevilla, Seville), <u>Javier Ruiz-Martínez</u> (Hospital Universitario Donostia, Instituto de Investigación Sanitaria Biodonostia, San Sebastián), <u>Clara Ruz</u> (Centro de Investigacion Biomedica, Universidad de Granada, Granada), <u>Antonio Sanchez Rodríguez</u> (Hospital Universitario Marqués de Valdecilla-IDIVAL, Santander), <u>María Sierra</u> (Hospital Universitario Marqués de Valdecilla-IDIVAL, Santander), <u>Esther Suarez-Sanmartin</u> (Hospital Universitario Central de Asturias, Oviedo), <u>Cesar Tabernero</u> (Hospital General de Segovia, Segovia), <u>Juan Pablo Tartari</u> (Fundació Docéncia i Recerca Mútua de Terrassa and Movement Disorders Unit, Department of Neurology, University Hospital Mutua de Terrassa, Terrassa, Barcelona), <u>Cristina Tejera-Parrado</u> (Instituto de Biomedicina de Sevilla (IBiS), Hospital Universitario Virgen del Rocío/CSIC/Universidad de Sevilla, Seville), <u>Eduard Tolosa</u> (Hospital Clinic Barcelona), <u>Francesc Valldeoriola</u> (Hospital Clinic Barcelona), <u>Laura Vargas-González</u> (Instituto de Biomedicina de Sevilla (IBiS), Hospital Universitario Virgen del Rocío/CSIC/Universidad de Sevilla, Seville), <u>Lydia Vela</u> (Department of Neurology, Hospital Universitario Fundación Alcorcón, Madrid), <u>Francisco Vives</u> (Centro de Investigation Biomedica, Universidad de Granada, Granada).

<u>**Austria:**</u>

<u>Alexander Zimprich</u> (Department of Neurology, Medical University of Vienna, Austria)

<u>**Norway:**</u>

<u>Lasse Pihlstrom</u> (Department of Neurology, Oslo University Hospital, Oslo, Norway), <u>Mathias Toft</u> (Department of Neurology and Institute of Clinical Medicine, Oslo University Hospital, Oslo, Norway)

<u>**Estonia:**</u>

<u>Pille Taba</u> (Department of Neurology and Neurosurgery, University of Tartu, Tartu, Estonia)

<u>**Australia:**</u>

<u>Sulev Koks</u> (Centre for Molecular Medicine and Innovative Therapeutics, Murdoch University, Murdoch, 6150, Perth, Western Australia; The Perron Institute for Neurological and Translational Science, Nedlands, 6009, Perth, Western Australia)

<u>**Israel:**</u>

<u>Sharon Hassin-Baer</u> (The Movement Disorders Institute, Department of Neurology and Sagol Neuroscience Center, Chaim Sheba Medical Center, Tel-Hashomer, 5262101, Ramat Gan, Israel, Sackler Faculty of Medicine, Tel Aviv University, Tel Aviv, Israel)

<u>**Finland:**</u>

<u>Kari Majamaa</u> (Institute of Clinical Medicine, Department of Neurology, University of Oulu, Oulu, Finland; Department of Neurology and Medical Research Center, Oulu University Hospital, Oulu, Finland), <u>Ari Siitonen</u> (Institute of Clinical Medicine, Department of Neurology, University of Oulu, Oulu, Finland; Department of Neurology and Medical Research Center, Oulu University Hospital, Oulu, Finland), <u>Pentti Tienari</u> (Clinical Neurosciences, Neurology, University of Helsinki, Helsinki, Finland, Helsinki University Hospital, Helsinki, Finland)

<u>**Nigeria:**</u>

<u>Njideka U. Okubadejo</u> (University of Lagos, Lagos State, Nigeria), <u>Oluwadamilola O. Ojo</u> (University of Lagos, Lagos State, Nigeria),

<u>**Kazakhstan:**</u>

<u>Coordinator - Rauan Kaiyrzhanov</u> (Department of Molecular Neuroscience, UCL Institute of Neurology, London, UK), <u>Chingiz Shashkin</u> (Kazakh National Medical University named after Asfendiyarov, Almaty, Kazakhstan), <u>Nazira Zharkinbekova</u> (South Kazakhstan Medical Academy, Shymkent, Kazakhstan), <u>Vadim Akhmetzhanov</u> (Astana Medical University, Astana Kazakhstan), <u>Gulnaz Kaishybayeva</u> (Scientific and practical center “Institute of neurology named after Smagul Kaishibayev”, Almaty, Kazakhstan), <u>Altynay Karimova</u> (Scientific and practical center “Institute of neurology named after Smagul Kaishibayev”, Almaty, Kazakhstan), Talgat Khaibullin (Semey Medical University, Semey, Kazakhstan).

<u>**Ireland:**</u>

<u>Timothy L. Lynch</u> (The Dublin Neurological Institute at the Mater Misericordiae University Hospital, Dublin, Ireland & School of Medicine and Medical Science, University College Dublin, Dublin, Ireland).

## Data access

BMJ and AJN had full access to all the data in the study and take responsibility for the integrity of the data and the accuracy of the data analysis

